# The Gini Coefficient as a useful measure of malaria inequality among populations

**DOI:** 10.1101/2020.09.19.20197939

**Authors:** Jonathan Abeles, David J Conway

## Abstract

**BACKGROUND:** Understanding inequality in infectious disease burden requires clear and unbiased indicators. The Gini coefficient, conventionally used as a macroeconomic descriptor of inequality, is potentially useful to quantify epidemiological heterogeneity. With a potential range from 0 (all populations equal) to 1 (populations having maximal differences), this coefficient is used here to show the extent and persistence of inequality of malaria infection burden at a wide variety of population levels.

**METHODS:** We first applied the Gini coefficient to quantify variation among WHO world regions for malaria and other major global health problems. Malaria heterogeneity was then measured among countries within the geographical sub-region where burden is greatest, among the major administrative divisions in several of these countries, and among selected local communities. Data were analysed from previous research studies, national surveys, and global reports, and Gini coefficients were calculated together with confidence intervals using bootstrap resampling methods.

**RESULTS:** Malaria showed a very high level of inequality among the world regions (Gini coefficient, *G* = 0.77, 95% CI 0.66-0.81), more extreme than for any of the other major global health challenges compared at this level. Within the most highly endemic geographical sub-region, there was substantial inequality in estimated malaria incidence among countries of West Africa, which did not decrease between 2010 (*G* = 0.28, 95% CI 0.19-0.36) and 2018 (*G* = 0.31, 0.22-0.39). There was a high level of sub-national variation in prevalence among states within Nigeria (*G* = 0.30, 95% CI 0.26-0.35), but more moderate variation within Ghana (*G* = 0.18, 95% CI 0.12-0.25) and Sierra Leone (*G* = 0.17, 95% CI 0.12-0.22). There was also significant inequality in prevalence among local village communities, generally more marked during dry seasons when there was lower mean prevalence. The Gini coefficient correlated strongly with the Coefficient of Variation which has no finite range.

**CONCLUSIONS:** The Gini coefficient is a useful descriptor of epidemiological inequality at all population levels, with confidence intervals and interpretable bounds. Wider use of the coefficient would give broader understanding of malaria heterogeneity revealed by multiple types of studies, surveys and reports, providing more accessible insight from available data.

## BACKGROUND

Describing levels of inequality of disease burden among populations is vital for epidemiology and global health, to highlight those who are affected disproportionately, and better target control interventions [1]. Most infectious disease and epidemiological reports do not give clear quantitative overviews on inequality, and the topic has been noted as requiring more attention in pursuit of the United Nations Sustainable Development Goals (UNSDGs) [2]. The particular UNSDG focusing on health includes an aim to end malaria as a public health problem by the year 2030 [3], encouraging control efforts, surveillance, and estimates of the situation through the World Health Organization (WHO) annual World Malaria Reports as well as national Malaria Indicator Surveys [4].

The Gini coefficient, an index widely used to describe income inequality, has been utilised previously to analyse general global health inequality [5, 6], sub-national differences in mortality [7], and for ecological studies [8], but only rarely for specific infectious diseases [9, 10]. Disease burden variation among populations is more commonly presented using general measures of dispersion such as interquartile range, standard deviation, and sometimes the coefficient of variation (a scale-invariant coefficient obtained by dividing the standard deviation by the mean) [11]. However, these typical descriptors do not easily facilitate comparisons, whereas a benefit in using the Gini coefficient of inequality is that it has a standardised range from 0 (all populations equal) to 1 (populations having maximal differences) so that levels of inequality can be benchmarked.

Here the Gini coefficient is applied to illustrate inequality in malaria at several population levels, global, regional, sub-national and local. This shows that global inequity is higher than for other diseases, and is not decreasing in the areas that are most highly affected, while there are significant differences among countries in levels of sub-national inequity, and local as well as seasonal variation.

## METHODS

### Gini coefficient of inequality

The Gini coefficient is a measure of variation among different populations or groups, either of a positive resource or of an undesired burden, that is derived from the Lorenz curve of inequality [12]. In macroeconomics, it compares the proportion of ‘wealth’ owned by each single sub-population or group and describes the resulting inequality when considering the total. Applied to malaria or other diseases, it can be focused on comparing data at any level, for example among geographical regions globally or among local communities within a defined area. In such analysis, the Lorenz curve estimates how the distribution of disease burden (or other relevant measure such as infection prevalence) deviates from a theoretical line of perfect equality, and this deviation is summarised in the Gini coefficient. A coefficient of 1 is ‘perfectly unequal’, whereas a zero value represents ‘perfectly equal’ distribution.

The Gini coefficient (*G*) is estimated by comparing the values of the relevant indicator (such as prevalence) among all populations, and calculating all pairwise differences among them, using the following formula:

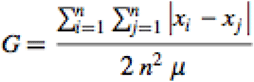

where *x*_*i*_ is the value for each individual population *i*, and *x*_*j*_ is the value for each of the individual populations *j* with which it is compared pairwise, there are *n* populations and *µ* is the mean value across all populations.

Gini coefficients were calculated in this study using STATA Version 15.3, with replicate analyses performed using Microsoft Excel and R version 3.6.3 to ensure consistency. Confidence intervals around the Gini coefficient were calculated using bootstrap resampling [13]. Equal bootstrap samples of size *n* are repeatedly drawn by sampling, and data are replaced after each sample. Bootstrap confidence intervals were calculated using R version 3.6.3, using command scripts as detailed in Supplementary Text S1, with resampling of *k* = 500 replicates as there is no significant benefit in using higher values of *k* in this context [14].

### Global and regional estimates for malaria and other public health challenges

Variation in disease burden across the different WHO world regions (African Region, Region of the Americas, South-East Asia Region, European Region, Eastern Mediterranean Region, and Western Pacific Region) was first analysed using estimates of numbers of cases in the most recent WHO annual report or fact sheet for each global health problem. Malaria was analysed [4], as well as tuberculosis [15], HIV/AIDS [16], and Hepatitis C infection [17] representing major global infectious diseases, and the four groups of non-communicable diseases with highest overall mortality (cancer, respiratory disease, cardiovascular disease, and diabetes) [18-20]. Data were expressed as estimated number per 100,000 of the population in each region before calculating Gini coefficients (Supplementary Table S1). Malaria estimates were also presented as a percentage proportion of population at risk yearly between 2010 and 2018 (Supplementary Table S2).

The Gini coefficient was also calculated for variation across 16 West African countries using World Malaria Report estimates of numbers of cases for each year between 2010 and 2018 (Supplementary Table S3) [4], based on estimated numbers of cases in proportion to the number of people in areas of risk.

### Malaria data at national, sub-national and local levels

Recent national Malaria Indicator Surveys (MIS) in four West African countries (Nigeria, Ghana, Sierra Leone and Burkina Faso) were analysed to compare the levels of sub-national variation in heavily affected countries [21-24]. Data were analysed from surveys of infection prevalence in under 5 year old children in 35 States in Nigeria (excluding one state that did not have sufficient data) [21], 10 Regions as previously defined in Ghana [22], 13 Regions in Burkina Faso [23], and 14 Districts in Sierra Leone [24]. Sub-national prevalence measurements from the MIS data analysed are tabulated in Supplementary Table S4.

To illustrate local variation in malaria infection prevalence and investigate temporal variation in highly endemic communities, data from The Garki Project were examined, given the availability of data from multiple cross-sectional surveys of multiple local village communities in an area of high prevalence in northern Nigeria in the early 1970s [25], from the archived database (http://garkiproject.nd.edu/). This analysis focused on an 18-month pre-intervention phase of the study during which eight successive parasitological cross-sectional surveys were conducted approximately every 10 weeks in each of 16 villages (data on a further six villages were not analysed as they were not surveyed at all eight rounds). Presence of *P. falciparum* was determined by microscopy and village-specific prevalence of *P. falciparum* at each survey is shown in Supplementary Table S5, after extraction from the archived project database. Data from a more recent study from The Gambia were also analysed for comparison, incorporating 20 villages with similar malaria seasonality [26], with variation in percent prevalence of *P. falciparum* determined by microscopy compared among villages in wet and dry seasons. Gini coefficient estimation from these and other previous data was performed using STATA version 15.3 with bootstrap confidence intervals calculated using R version 3.6.3.

## RESULTS

### Global variation in major health problems

Across the six WHO world regions, inequality was greater for each of the infectious diseases compared to the major non-communicable diseases (Figure 1). There was most extreme inequality in malaria burden (Gini coefficient, *G* = 0.77, 95% CI 0.66-0.81), which was significantly higher than for each of the other diseases as demonstrated by bootstrap confidence intervals (Figure 1 and Supplementary Table S1).

**Figure 1.**
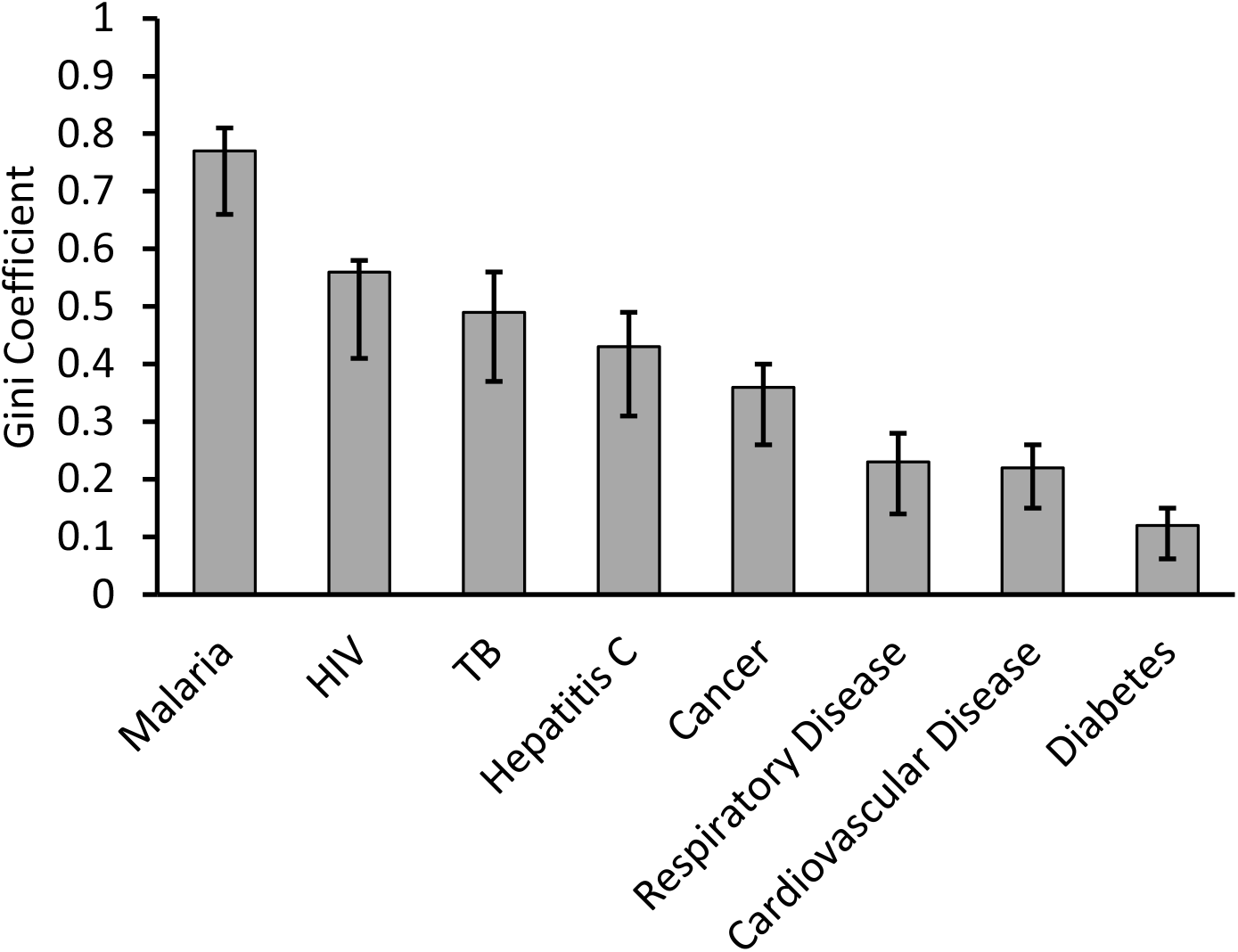
Exceptionally high global inequality of malaria burden compared with other major public health indices. Gini coefficients of inequality of disease burdens among the six major regions of the world were calculated for eight major public health problems. The world regions are Africa, The Americas, South East Asia, Europe, Western Pacific, and Eastern Mediterranean as defined by the World Health Organization (WHO). Analyses are based on data or estimates extracted from the most recent fact sheets or world reports, or Global Health Estimates by WHO (listed in Supplementary Table 1). Malaria, HIV, and tuberculosis (TB) estimates represent new infections in 2018 [4, 15, 16], while Hepatitis C estimates represent new infections in 2015 [17]. For the four non-communicable diseases with highest mortality, different types of estimates are used as examples: diabetes estimates were based on prevalence in adults in 2014 [18], cancer estimates refer to new cases in the year of 2018 [19], while estimates for cardiovascular and respiratory disease refer to attributable deaths in 2016 [20]. The inequality of malaria burden was higher than for the other indices, as shown by the Gini coefficient estimates (with 95% bootstrap confidence intervals).

Removing the Europe WHO region from the malaria calculation (which had no reported cases in 2018) did not greatly reduce the index of inequality (*G* = 0.73, 95% CI 0.59-0.77). Removing the African WHO region (containing approximately 90% of all malaria cases), showed residual inequality among remaining regions to be much lower but still substantial (*G* = 0.40, 95% CI = 0.26-0.54), reflecting that most other global cases are in Asia or the Western Pacific. There was no decline in levels of global inequality between 2010 and 2018 based on data estimates from the World Malaria Report (*G* values for each year remained between 0.76 and 0.78, Supplementary Table S2).

### Variation in malaria within West Africa

As the African region has the majority of the malaria burden, and more than half of the cases are estimated to occur in West Africa, we analysed the inequality among the 16 countries that constitute The West African sub-region according to the UN definition. This shows the variation in estimated malaria burden as a proportion of overall populations among countries in West Africa between 2010 and 2018, and the persistent inequality is revealed by the Gini coefficient (Figure 2 and Supplementary Table S3). Although there have been moderate reductions in malaria overall, and notable reductions in a few countries, the average burden is still high and variation among countries persists (Figure 2A). Accordingly, the Gini Coefficient of inequality remained high, between 0.27 and 0.32 in each year (Figure 2B). In 2018, the final year estimated, there was still marked inequality in malaria burden among countries in West Africa (*G* = 0.31, 95% CI 0.22-0.39) and no indication of this having reduced from 2010 onwards.

**Figure 2.**
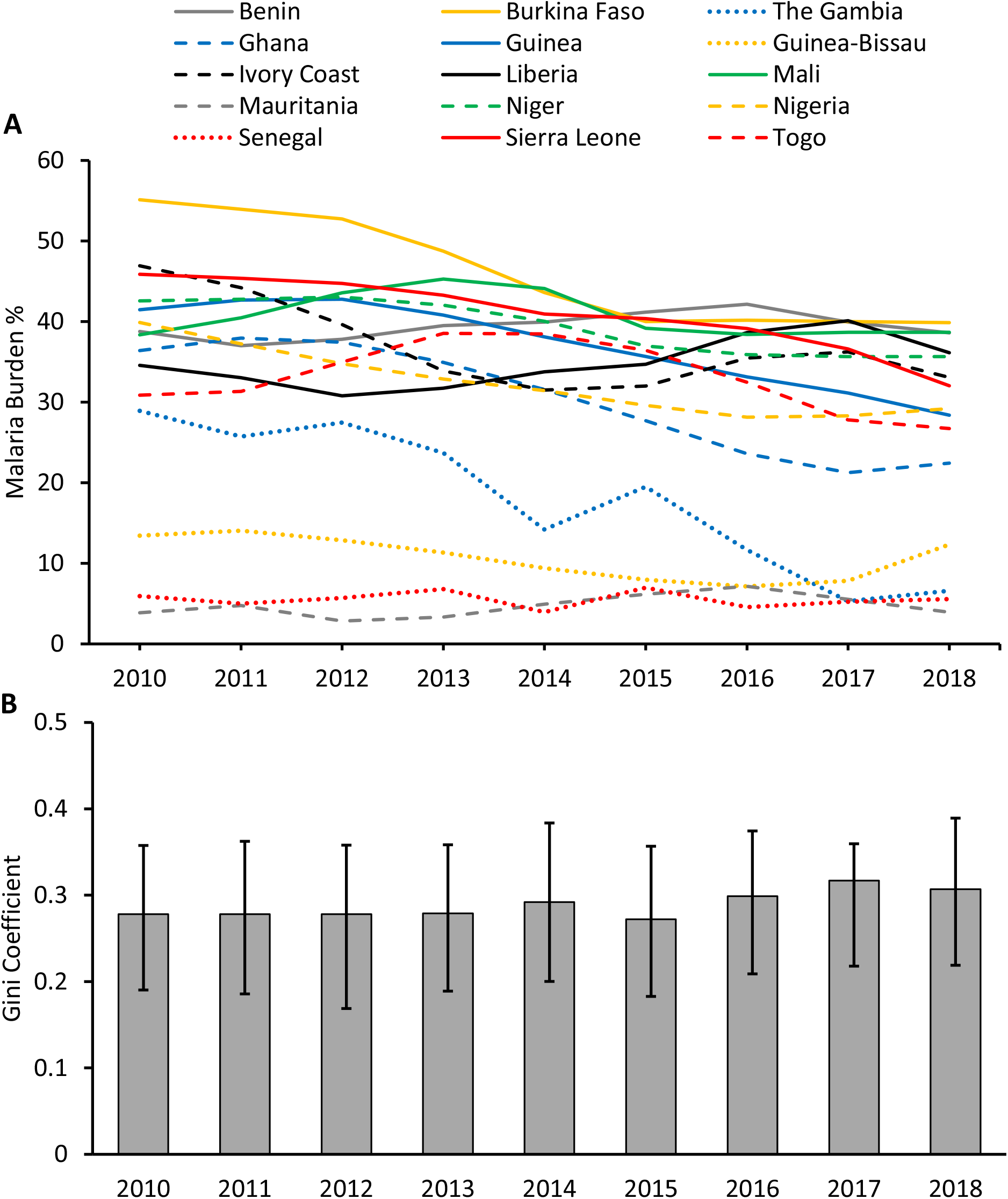
Inequality of malaria burden among countries in West Africa has not reduced between 2010 and 2018. **A**. Malaria burden in individual West African countries was calculated as the estimated annual number of cases divided by population at risk. Estimated number of cases were as presented in the WHO World Malaria Report annexes [4], divided by the estimated population at risk (Supplementary Table 3). Data are plotted for all countries except Cape Verde for which numbers of cases were at or close to zero in each year. **B**. The Gini coefficient estimates (with 95% confidence intervals) show no decreases in inequality of malaria burden among the countries over time, values of the coefficient being moderately high and remaining between 0.27 and 0.31 in all years (with overlapping 95% confidence intervals).

### Sub-national variation in malaria within high burden countries in West Africa

Sub-national variation within four of the highest burden countries was analysed using Malaria Indicator Survey data of malaria infection prevalence in children under 5 years of age, with surveys between the years of 2014 and 2016 allowing prevalence to be analysed for each of the principal formal administrative divisions within each country (States in Nigeria, Regions in Ghana and Burkina Faso, Districts in Sierra Leone) (Figure 3 and Supplementary Table S4). Sierra Leone had the highest mean prevalence, but the highest level of sub-national inequality in malaria parasite prevalence was seen in Nigeria (*G* = 0.30, 95% CI 0.26-0.35), followed by Burkina Faso (*G* = 0.25, 95% CI 0.19-0.29), with more moderate sub-national inequality within Ghana (*G* = 0.18, 95% CI 0.12-0.25) and Sierra Leone (*G* = 0.17, 95% CI 0.12-0.22).

**Figure 3.**
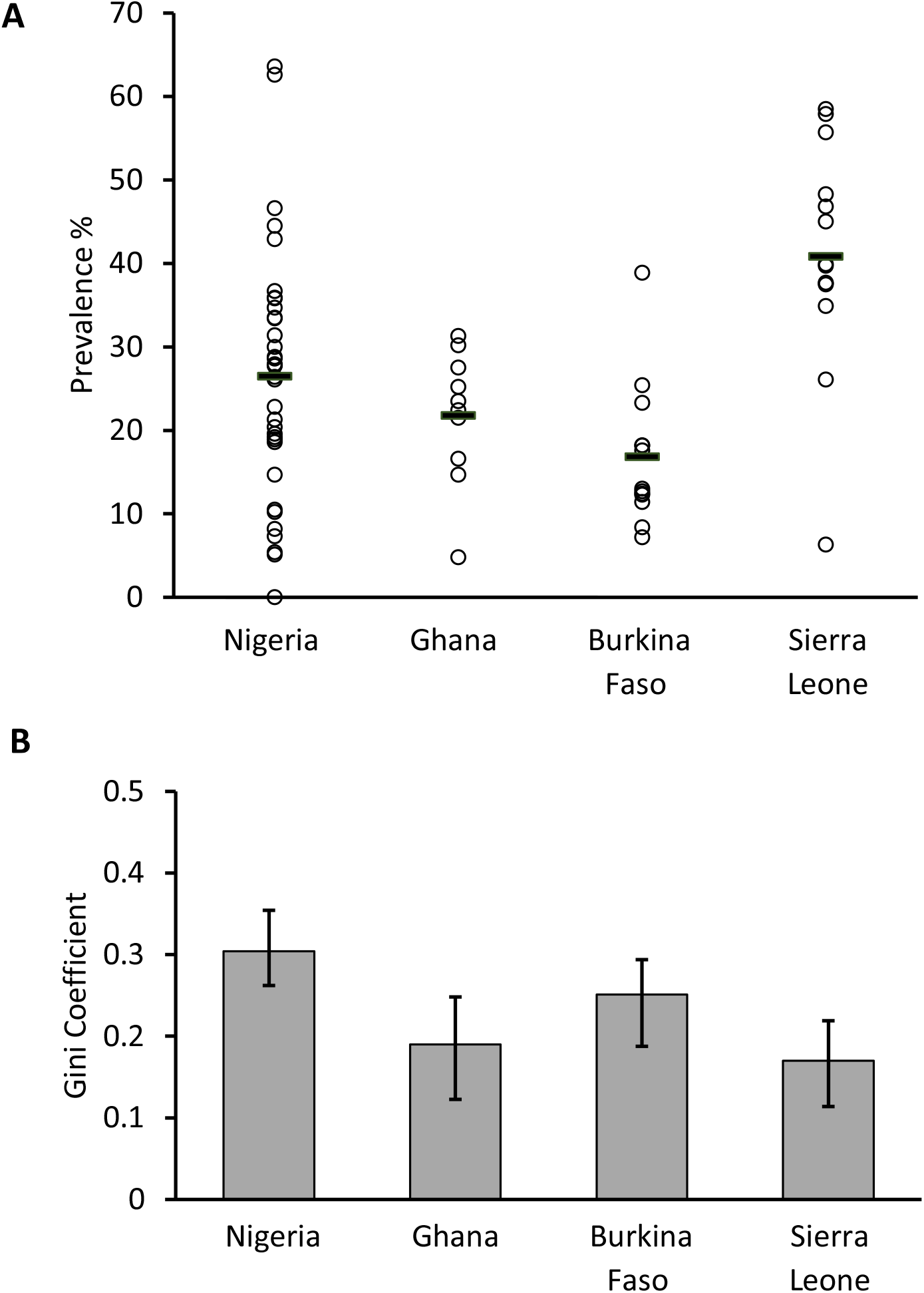
Inequality of malaria prevalence among major administrative regions within four high burden countries in West Africa. **A**. Each point represents the community prevalence of slide-positive malaria infection in children 6-59 months of age in the major administrative geographical regions within each of four countries as reported their most recent malaria indicator surveys (MIS). Data are analysed for 35 States in Nigeria from the 2015 MIS (having excluded one that did not have sufficient data) [21], 10 Regions within Ghana from the 2016 MIS [22], 13 Regions in Burkina Faso from the 2017 MIS [23], and 14 Districts in Sierra Leone from the 2016 MIS [24], as presented in Supplementary Table 4. **B**. The within-country Gini coefficient of inequality was highest for Nigeria (*G* = 0.30, 95% CI 0.26-0.35), indicating a similar amount of variation within that country as exists among all 16 countries of West Africa. This was significantly greater variation than within Ghana or Sierra Leone.

### Seasonal malaria heterogeneity among villages in a highly endemic area

Local variation was investigated among villages in the Garki Project, a large study conducted in northern Nigeria in the early 1070s [25]. The *P. falciparum* prevalence was compared among 16 villages for which there were data at eight different survey timepoints (each survey separated by approximately 10 weeks) during the pre-intervention phase of the project (Figure 4A and Supplementary Table S5). An overall seasonal peak of malaria prevalence is evident which corresponds to the late wet season and immediate post-wet season (surveys 5 and 6), while lower prevalence is seen during the annual dry seasons (surveys 2 and 3 for one year, and surveys 7 and 8 for the following year). The Gini coefficients are highest at survey timepoints 2, 3, and 7, which coincide with the dry seasons (Figure 4B). This demonstrates the ability of the Gini coefficient to track local variation in epidemiology, including temporal changes and effects of seasonality. To test for consistency in variation among villages across years, the rank order of prevalence in villages at survey 2 and survey 7 (representing similar points in consecutive dry seasons) was tested and shown to be significantly correlated (Spearman’s rho = 0.55, P=0.028). This indicates that a significant component of the inter-village variation was maintained for at least a year.

**Figure 4.**
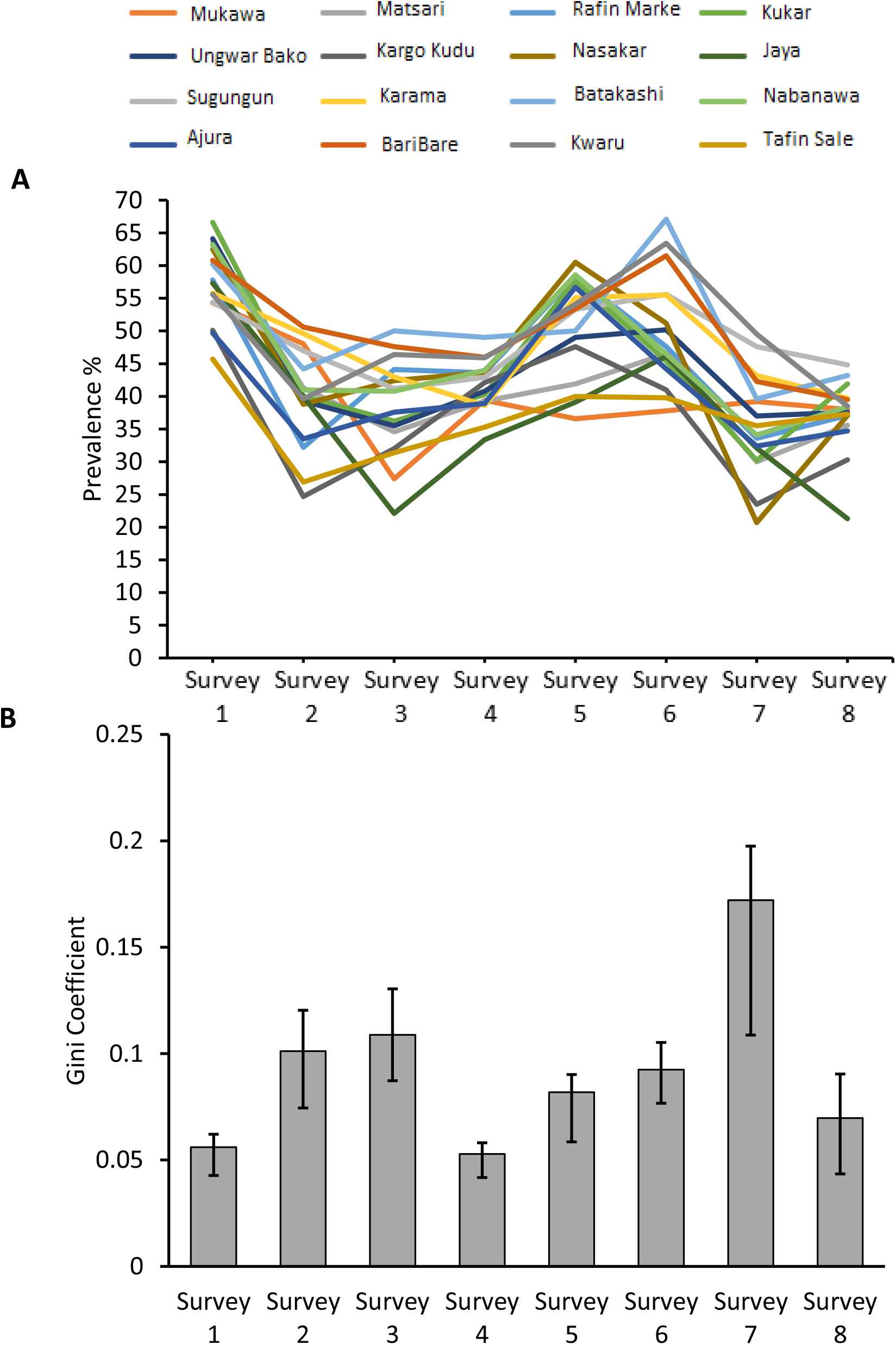
Local inter-village heterogeneity in malaria prevalence varies over time and shows seasonality captured by the Gini coefficient. Temporal variation is analysed in data from successive community surveys during the pre-intervention phase of The Garki Project, a classic epidemiological study previously conducted in a rural district in northern Nigeria. **(A)** Malaria parasite slide positive prevalence data were obtained from 8 cross-sectional surveys conducted 10 weeks apart (spanning 70 weeks in total). Annual peak of infection prevalence was seen during and immediately following the rainy season (survey 5 and 6), and lower prevalence is evident during and immediately following the dry season (surveys 2 and 3 for one year, surveys 7 and 8 for the following year). Data are shown for the 16 villages that had surveys at all 8 timepoints (data from a total of 5797 participants were included). Prevalence data were extracted from The Garki Project archived database (http://garkiproject.nd.edu/) and are shown in Supplementary Table S5. **(B)** Gini coefficients representing the extent of inter-village heterogeneity in malaria in each of the surveys with bootstrap confidence intervals. Overall variation among the villages was moderately low, with a peak G = 0.17, with significant seasonal variation in Gini coefficients as most inequality occurred in the dry season when average prevalence was lower.

Data were then analysed from a study of 20 villages in The Gambia [26], where malaria endemicity is lower and surveys were conducted shortly after malaria had declined significantly throughout the country [27, 28]. Comparing across all villages surveyed, heterogeneity was higher in the dry season (*G* = 0.55) than the wet season (*G* = 0.40). Focusing on the eastern part of The Gambia where malaria prevalence is highest, there was greater contrast in variation among villages in the dry season compared to the wet season (Supplementary Figure S1). Variation among villages was generally higher than seen in the data from The Garki Project in which the infection prevalence was higher at all times of the year.

### Comparison to the Coefficient of Variation

There is a strong correlation between the values of the Gini Coefficient presented here, and the values of the Coefficient of Variation (CV, standard deviation divided by the mean) calculated for each of the same datasets (Spearman’s rho = 0.982, P < 10^−4^). Gini coefficient values ranged from 0.05 to 0.77, while CV values ranged from 9 % to 198 %, with a strong correlation over the whole range (Figure 5). As the Gini coefficient is bounded between 0 and 1 and has bootstrap confidence intervals (not plotted in Figure 5), it offers advantages for interpretation compared to the use of an unbounded CV index.

**Figure 5.**
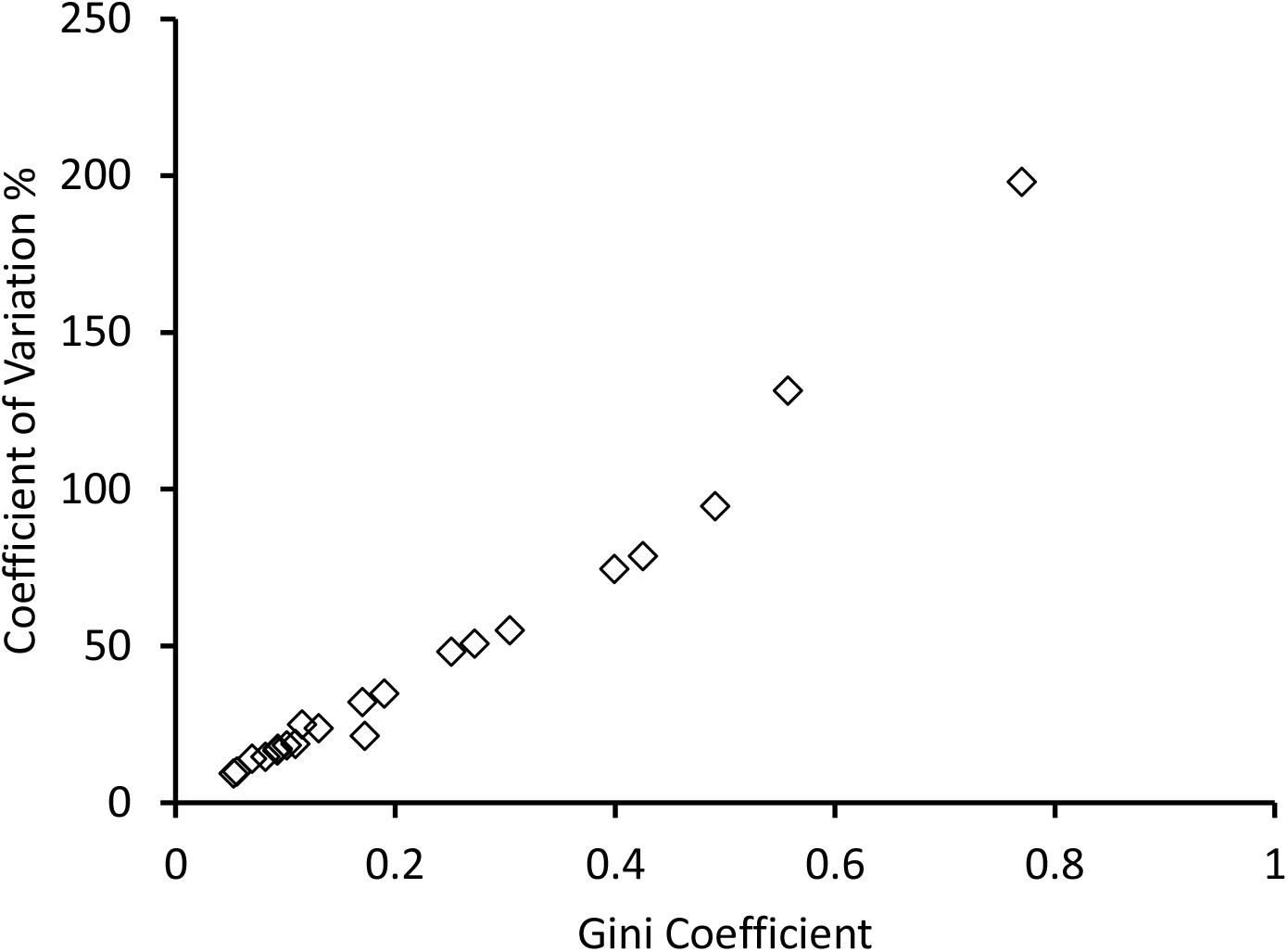
Gini Coefficient compared to Coefficient of Variation for inter-population comparisons. This shows strong correlation between the Gini coefficient and the standard coefficient of variation (standard deviation divided by the mean) illustrating the Gini coefficient as a robust general statistical descriptor of variation. This is alongside its interpretive utility as being bounded between 0 and 1 (whereas the Coefficient of Variation is an unbounded proportional value that does not offer an intuitive scale for epidemiological comparisons). There were 21 values compared for each index here, representing data from different analyses presented in this paper: eight for various diseases among world health regions (from Figure 1), one for malaria across the countries of West Africa (Figure 2), four of sub-national variation in malaria prevalence in selected West African countries (Figure 3), and eight sequential surveys across villages in The Garki Project (Figure 4).

## DISCUSSION

Inequality in malaria burden among populations is effectively summarised into a single index using the Gini coefficient, as shown here. Among leading global infectious and non-communicable public health problems, malaria shows the highest amount of inequality among different world regions, with a Gini coefficient of 0.77 being closer to the theoretically maximum possible value of 1.0 than to zero which would indicate equitable distribution. This coefficient has not been reduced in recent years, so there clearly needs to be increased effort to reducing the malaria burden in the most highly affected African region, while sustaining recent reductions of malaria elsewhere. This global need is already qualitatively clear [4], but the use of the Gini coefficient highlights the extreme situation for malaria in comparison to other diseases, and shows the measurability of inequality which is essential for future monitoring of progress.

Of equal importance, the Gini coefficient is also shown to be useful for summarising inequality at other population levels, from regional to local. Within West Africa, the sub-region with the highest overall malaria burden globally, the coefficient shows that malaria inequality among countries has not declined in recent years, reflecting that relative reductions in malaria burden have not been particularly great in the countries with most malaria. Moreover, the amount of sub-national inequality within four high burden countries in West Africa is also shown to be significantly variable. For example, there is more inequality in the infection prevalence among different states in Nigeria than among the major administrative areas within Ghana or Sierra Leone, analysing data from national Malaria Indicator Surveys that employ broadly comparable survey methods. The causes of such sub-national inequality will be complex and require more research attention for future malaria control.

The Gini coefficient is sensitive to village-level, area-level, and seasonal variation, as illustrated here by re-analyses of research survey data from studies previously conducted in different parts of West Africa. The coefficient has features that make it potentially a more useful descriptor of epidemiological heterogeneity than other summary indices. The Gini coefficient demonstrates a defined lower and upper boundary of 0 (perfect equality) and 1 (perfect inequality) while the coefficient of variation (CV) based on standard deviation does not. Although there is a strong correlation in their quantification of heterogeneity, the CV summarises variation in an unbounded range that can transcend 100%. The Gini coefficient is therefore more appropriate for use in the context of epidemiological studies and disease reports, to enable a more standardised quantitative interpretation of inequality.

While the Gini coefficient is a useful descriptor, limitations should be considered. Technically, although bootstrap resampling is a generally robust method of calculating confidence intervals for the Gini coefficient, it has been suggested that in small samples of uniform, normal, or lognormal distributions bootstrap confidence intervals may be calculated as too narrow [14, 29], and robustness of these intervals increases with larger numbers of sampled populations. Statistical methods have been developed to mitigate this issue by approximating the Lorenz curve of a log-normal distribution [30], and could be investigated in future to check the sensitivity of confidence interval estimations. Also, although this was not a particular issue with the data analysed here, Gini coefficients could be skewed by ‘small number bias’ if they were based on samples of populations with extremely low prevalence, essentially corresponding to sampling noise that gives a systematic bias towards an inflated Gini coefficient in such situations.

Epidemiologically, we note that the Gini coefficient needs to be recognised as a simple relative measure that does not present absolute differences, and different distributions of measurements may produce the same Gini coefficient. Demographic and socioeconomic differences, as well as ecological, genetic and geographical determinants all combine together [31, 32] and the effect of each is not explored or separately accounted for by the Gini coefficient. Clearly, the coefficient does not substitute for separate analyses of the epidemiological determinants, or for maps of disease distribution, where these are available or where they may be estimated [33, 34]. Instead, it should be applied alongside presentation of more detailed or qualitative data, and used to advocate focus on populations most affected and where control of malaria is most needed, aiming to reduce the extreme inequity that continues to prevail at multiple levels.

## CONCLUSION

The Gini coefficient is a useful index for descriptive epidemiology, particularly relevant for malaria which shows exceptional levels of global inequality compared to other diseases. As illustrated here, it is applicable to a wide variety of data sources to highlight the degree of inequality among world regions, or countries within a region, as well as sub-nationally or locally. We encourage its use more widely in the presentation of prevalence data, as it would be straightforward to interpret in publications including disease reports, national indicator surveys, and for a broad range of population-based research.

## Data Availability

Previous epidemiological data analysed here are tabulated in Supplementary Information, and all references given. The methods are fully given in the text and Supplementary Information.

## DECLARATIONS

### Ethics Approval and Consent to Participate

The Research Governance and Integrity Office of the London School of Hygiene and Tropical Medicine approved this analysis (reference 17018), and declared it not to require separate review by the Ethics Committee as it involved secondary analyses of data already in the public domain.

### Consent for publication

Not applicable.

### Authors’ contributions

JA contributed to study design, performed analyses and co-wrote the manuscript. DJC conceived the study, contributed to study design, supervised analyses and co-wrote the manuscript. Both authors reviewed and approved the final manuscript.

## Acknowledgements

We are grateful to colleagues who have discussed or commented on the analyses.

